# Early impact of the COVID-19 pandemic and social distancing measures on routine childhood vaccinations in England, January to April 2020

**DOI:** 10.1101/2020.05.07.20094557

**Authors:** Helen I McDonald, Elise Tessier, Joanne M White, Matthew Woodruff, Charlotte Knowles, Chris Bates, John Parry, Jemma L Walker, J. Anthony Scott, Liam Smeeth, Joanne Yarwood, Mary Ramsay, Michael Edelstein

**Affiliations:** NIHR Health Protection Research Unit (HPRU) in Immunisation; Immunisation and Countermeasures Division, Public Health England, Colindale, United Kingdom.; Statistics, Modelling and Economics Department, Public Health England, Colindale, United Kingdom.; Department of Infectious Disease Epidemiology, London School of Hygiene & Tropical Medicine, London, United Kingdom.; TPP (Leeds) Ltd, Leeds, United Kingdom

**Keywords:** COVID-19, pandemics, Measles-Mumps-Rubella vaccine, DTaP-IPV-Hib-HBV vaccine

## Abstract

Electronic health records were used to assess the early impact of COVID-19 on routine childhood vaccination in England to 26 April 2020.

MMR vaccination counts fell from February 2020, and in the three weeks after introduction of social distancing measures were 19.8% lower (95% CI −20.7 to −18.9%) than the same period in 2019, before improving in mid-April. A gradual decline in hexavalent vaccination counts throughout 2020 was not accentuated on introduction of social distancing.

---

Electronic patient records from primary care were analysed to investigate changes in delivery of the routine childhood vaccination programme in England during the COVID-19 outbreak to 26 April 2020.

## Near real-time records of routine childhood vaccinations from primary care

Two key milestones in the routine childhood immunisation programme delivered in primary care are first universal vaccinations at 8 weeks old, which include hexavalent vaccine (against diphtheria, tetanus, pertussis, polio, *Haemophilus influenzae* type b and hepatitis B), and vaccinations at one year old, which include the first dose of MMR vaccine (against measles, mumps and rubella).(1)

Aggregated weekly counts of first hexavalent vaccinations delivered to infants under 6 months, and first MMR vaccinations delivered to children aged 12 to 18 months, were provided from TPP SystmOne for the first 17 weeks of 2019 and 2020. SystmOne is a software system which provides electronic patient records for over 2600 primary care practices in the UK and over 35 child health providers. Data were anonymous throughout, having been extracted as aggregated weekly counts grouped by local Clinical Commissioning Group for population level data checks.

To minimise changes in denominator, only providers active in SystmOne since 2018 contributed to the dataset. The majority of vaccinations entered into SystmOne are entered by general practices in real time (over 70% in 2019). However, vaccinations delivered in general practices which use other patient record software may be recorded at a delay into the SystmOne integrated patient record by local Child Health systems. To distinguish changes in vaccine delivery from lags in data recording, only vaccinations recorded on the same day as they were delivered were included.

The dataset included 69,568 hexavalent doses delivered in 2019, and 67,116 in 2020; and 68,849 MMR doses delivered in 2019, and 66,301 in 2020.

## How did vaccination counts change during the COVID-19 outbreak?

Hexavalent vaccination counts followed a similar pattern in 2020 to 2019, varying week-by-week, (particularly low counts in week 1 are likely explained by the national bank holiday and school holidays) (**Figure 1**). MMR vaccination counts also followed a similar pattern in 2020 until week 11, when they fell sharply and remained low for several weeks before rising again in weeks 16 and 17.

**Figure 1:**
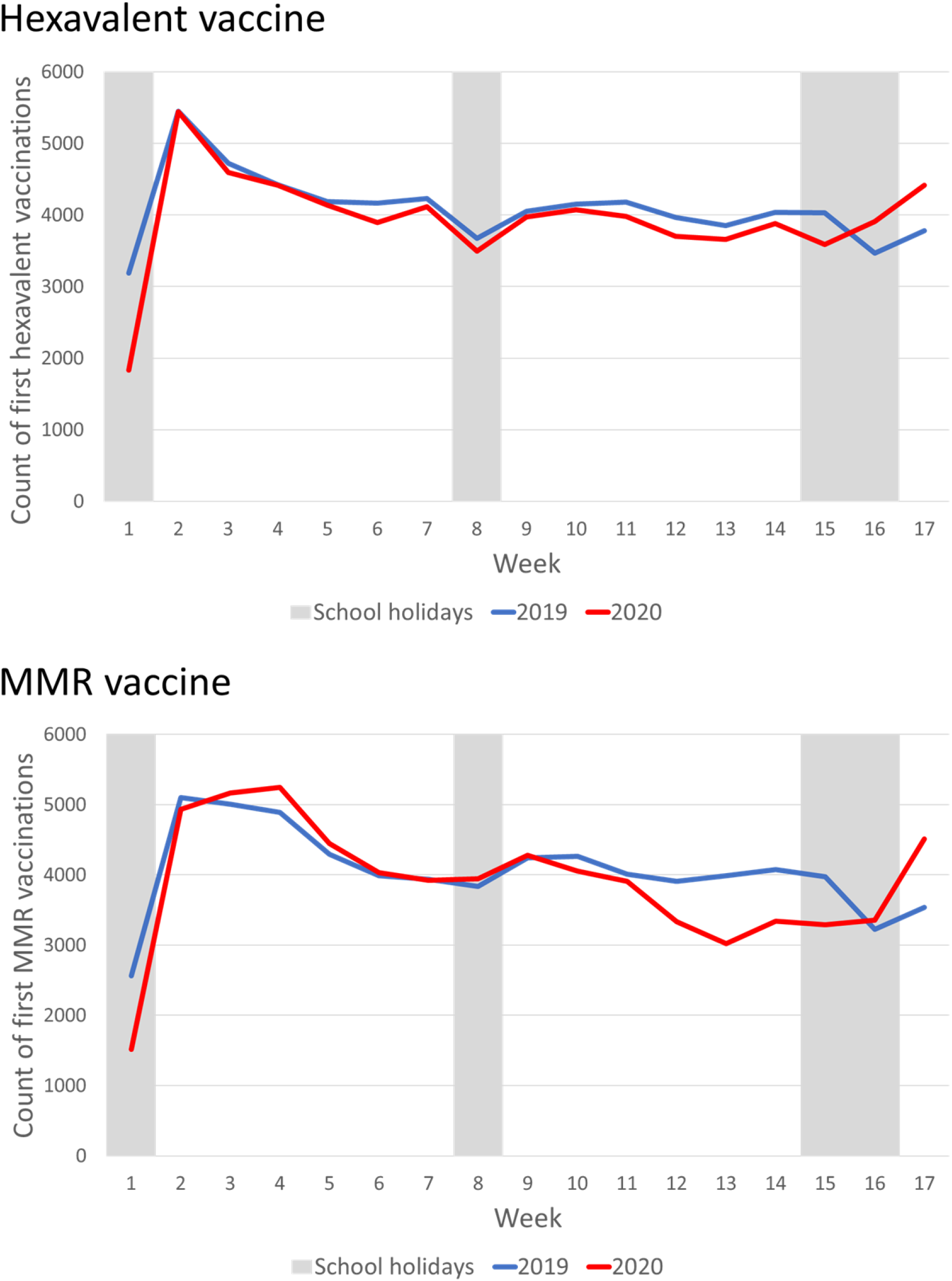
Hexavalent and MMR weekly vaccination counts in 2019 and 2020.

The percent change in vaccinations counts in 2020 compared to 2019 varied over the course of the COVID-19 outbreak (**Table 1**). Social distancing was a topic of public discussion in England from at least early March.(2) Prior to this, in weeks 1 to 9, hexavalent vaccination was 5.8% lower (95% CI −6.0 to −5.5%) and MMR vaccination 1.0% lower (95% CI −1.1 to −0.9%) in 2020 compared to 2019. On 12 March the UK government advised that anyone with a new continuous cough or a fever should self-isolate for seven days. In weeks 10 to 12 of 2020, hexavalent vaccination was 4.4% lower (95% CI −4.8 to −4.0%) and MMR vaccination 7.2% lower (95% CI −7.7 to −6.7%) than in 2019. Full social distancing measures were introduced on 20 March.(3) In the subsequent three weeks (weeks 13 to 15), hexavalent vaccination was 6.7% lower (95% CI −7.1 to −6.2%) and MMR vaccination 19.8% lower (95% CI −20.7 to −18.9%) than in 2019. In contrast, vaccination counts in 2020 were higher in weeks 16 and 17 of 2020 than for the same weeks in 2019, for both vaccines.

**Table 1:** Hexavalent and MMR vaccination counts in 2020 compared to 2019, by week.

| Week | Hexavalent 1 |  |  | MMR 1 |  |  |
| --- | --- | --- | --- | --- | --- | --- |
|  | 2019<br>n | 2020<br>n | Percent change<br>% (95% CI) | 2019<br>n | 2020<br>n | Percent change<br>% (95% CI) |
| 1 | 3,193 | 1,835 | -42.5 (-45.2 to -39.8) | 2,564 | 1,515 | -40.9 (-43.9 to -38.0) |
| 2 | 5,447 | 5,441 | -0.1 (-0.2 to 0.0) | 5,098 | 4,935 | -3.2 (-3.7 to -2.7) |
| 3 | 4,720 | 4,591 | -2.7 (-3.2 to -2.3) | 5,005 | 5,170 | 3.3 (2.8 to 3.8) |
| 4 | 4,426 | 4,418 | -0.2 (-0.3 to -0.1) | 4,893 | 5,246 | 7.2 (6.5 to 8.0) |
| 5 | 4,185 | 4,134 | -1.2 (-1.6 to -0.9) | 4,292 | 4,445 | 3.6 (3.0 to 4.1) |
| 6 | 4,168 | 3,895 | -6.5 (-7.4 to -5.7) | 3,988 | 4,029 | 1.0 (0.7 to 1.3) |
| 7 | 4,229 | 4,114 | -2.7 (-3.2 to -2.2) | 3,940 | 3,920 | -0.5 (-0.7 to 1.3) |
| 8 | 3,676 | 3,497 | -4.9 (-5.6 to -4.1) | 3,839 | 3,943 | 2.7 (2.1 to 3.2) |
| 9 | 4,051 | 3,974 | -1.9 (-2.3 to -1.5) | 4,241 | 4,278 | 0.9 (0.6 to 1.2) |
| 10 | 4,151 | 4,075 | -1.8 (-2.2 to -1.4) | 4,262 | 4,057 | -4.8 (-5.5 to -4.1) |
| 11 | 4,180 | 3,981 | -4.8 (-5.4 to -4.1) | 4,010 | 3,906 | -2.6 (-3.1 to -2.1) |
| 12 | 3,967 | 3,704 | -6.6 (-7.5 to -5.8) | 3,905 | 3,332 | -14.7 (-16.0 to -13.4) |
| 13 | 3,855 | 3,657 | -5.1 (-5.9 to -4.4) | 3,990 | 3,024 | -24.2 (-25.9 to -22.5) |
| 14 | 4,039 | 3,882 | -3.9 (-4.5 to -3.3) | 4,079 | 3,345 | -18.0 (-19.4 to -16.6) |
| 15 | 4,030 | 3,591 | -10.9 (-12.0 to -9.8) | 3,975 | 3,290 | -17.2 (-18.6 to -15.8) |
| 16 | 3,469 | 3,912 | 12.8 (11.7 to 13.9) | 3,227 | 3,356 | 4.0 (3.3 to 4.7) |
| 17 | 3,784 | 4,415 | 16.7 (15.5 to 17.9) | 3,541 | 4,510 | 27.4 (25.9 to 28.8) |
| **Total** | 69,568 | 67,116 | -3.5 (-3.7 to -3.4) | 65,341 | 61,832 | -3.7 (-3.8 to -3.6) |

Trends over time in the percent change of vaccination counts in 2020 compared to 2019 were modelled using Joinpoint regression (v4.8.0.0), which finds the best fit for points of change in trend.(4) For hexavalent vaccination, this suggested a general decrease in vaccination over 2020 compared to 2019, which did not accentuate on introduction of social distancing, but reversed in week 15, with a percent increase in weeks 16 and 17 of 2020 compared to 2019 (**Figure 2**). The percent change of MMR doses delivered in 2020 compared to 2019 was steady until week 9, but then decreased steeply to a low point of −24.2% (95% CI −25.9 to −22.5%) in week 13 before also reversing, with a percent increase in weeks 16 and 17 of 2020 compared to 2019 (**Figure 2**).

**Figure 2:**
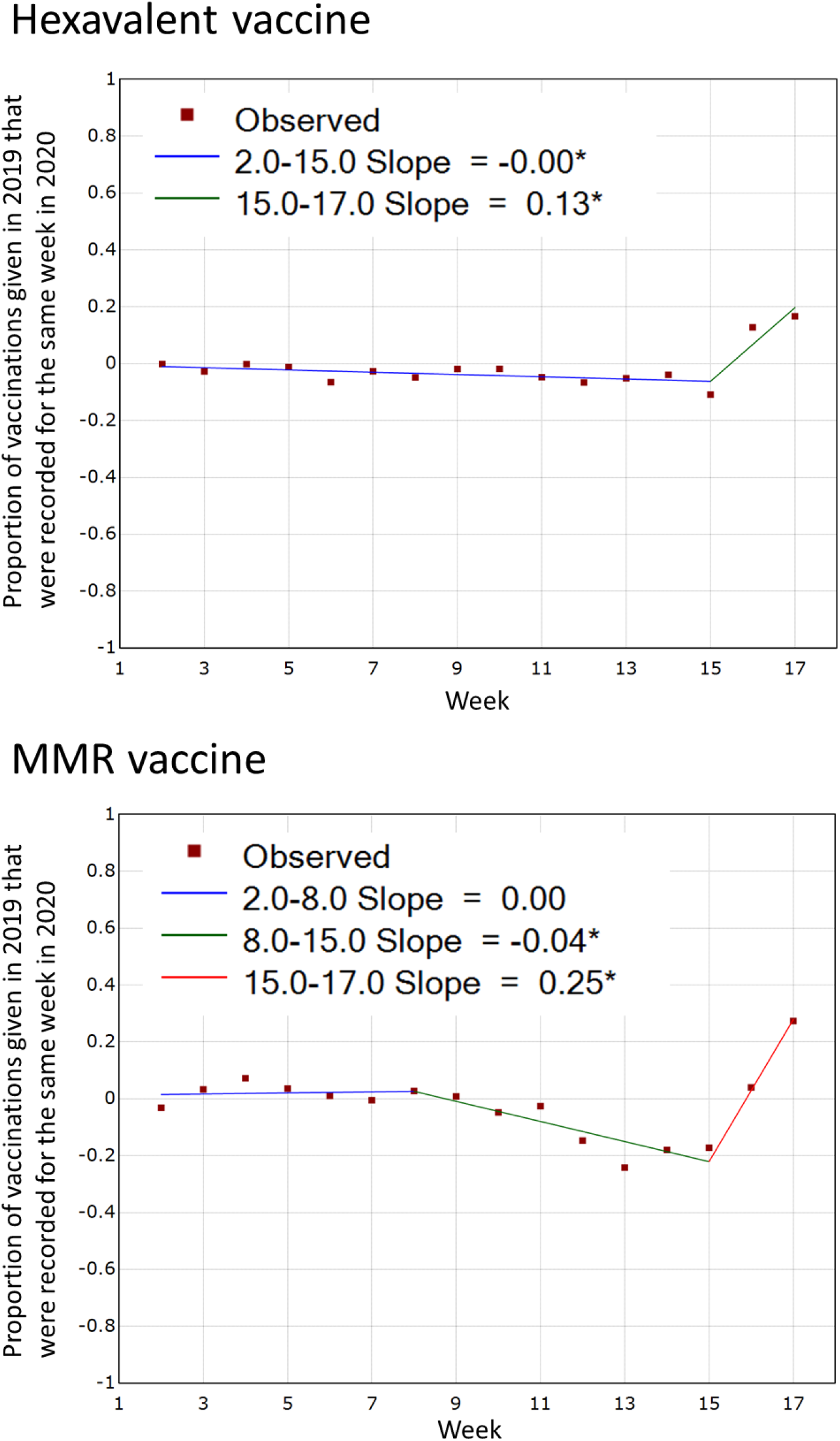
Percent change in hexavalent and MMR vaccination in 2020 compared to 2019, by week^*^. * Week 1 excluded, as the New Year’s Bank Holiday in week 1 fell on a Tuesday of in 2019 and Wednesday in 2020.

## How did changes in vaccination counts vary geographically?

In the three weeks following introduction of social distancing measures (weeks 13-15), the percent change in hexavalent vaccination in 2020 compared to 2019 varied by region, ranging from increases of 17.4% (95% CI 12.4 to 22.4%) in Cheshire and Merseyside to falls of more than 10% in Greater Manchester, London, the West Midlands and Yorkshire and the Humber. In contrast, MMR vaccination during weeks 13 to 15 was lower in 2020 than 2019 for all regions. The size of the percent decrease varied, with the greatest falls in London (−42.5%, 95%CI −48.2 to 36.9), Greater Manchester (−42.5%, 95%CI −55.4 to −29.7), and the West Midlands (−29.1%, 95%CI −34.8 to −23.3). By week 17 of 2020, the percent change in vaccination counts in 2020 compared to 2019 had improved in all regions, but only two regions had reached the cumulative vaccination count seen in week 17 of 2019.

## Discussion

MMR vaccination started falling in 2020 before introduction of social distancing measures. In the first three weeks of social distancing, MMR vaccination counts were 19.8% lower (95% CI −20.7 to −18.9%) than for the same period in 2019. There were signs of a recovery by mid-late April. There was a general decrease in hexavalent vaccinations delivered in 2020 compared to 2019, but no evidence of an increase in the rate of decline with the introduction of social distancing measures.

One plausible explanation is that COVID-19 messaging about staying home initially overwhelmed the message that the immunisation programme was to remain operating as usual. In England, this appears to have affected MMR vaccination more than primary infant vaccinations. The message to continue routine immunisation programmes became more visible when the Joint Committee for Immunisation and Vaccination published a statement on 17 April.(5) The relative increase in week 16 of 2020 compared to 2019 could partly be attributed to a low vaccination count in 2019 (coinciding with national holidays). However, a percent increase was also observed in week 17. This is promising for an early recovery in vaccination delivery, but will need monitoring to ensure it is sustained.

The greatest percent decrease in MMR vaccination was seen in London, which had a high early burden of COVID-19. However, decreases in MMR vaccination were also seen in regions with a low incidence of COVID-19 infection during this period, suggesting that the changes were not solely due to COVID-19 infection burden. SystmOne use varies regionally but it is unlikely that software system choice would result in vaccination counts being unrepresentative within regions.(6) Regional variation might also be explained by local communications, which may have mitigated the fall in some areas.

A limitation of this analysis is that changing counts of vaccinations delivered could be driven by numbers of eligible infants, rather than vaccine coverage. Decreasing birth rates may plausibly explain the gradually falling hexavalent vaccination counts, (7) and migration could play a role too, but these cannot explain the size and timing of the changes in MMR vaccination. Deferral of data entry could explain some decrease in real-time vaccination counts, but not the subsequent increase.

It is vital that routine childhood vaccinations continue, particularly for diseases such as measles for which a high coverage is required to prevent outbreaks.(5, 8) Decreased vaccine counts have also been reported in other high-income countries,(9, 10) and the Regional Office for Europe of the World Health Organization has advised that routine immunization services should continue to aim for high population immunity.(11) Countries will require immunisation recovery plans with innovative approaches to delivery that maintain social distancing requirements. Vaccinations usually delivered in schools and affected by school closures will also require catch-up programmes. Continuous and timely assessment of vaccine coverage will be required to respond to potentially volatile changes during the COVID-19 pandemic.

## Data Availability

The data were provided to Public Health England for the purposes of surveillance of routine childhood immunisation, and are not publicly available.

## Ethics approval

This analysis was conducted as part of public health usual practice, and was not conducted for research. Ethics approval was therefore not sought.

